# Evaluation of the Pilot Wastewater Surveillance for SARS-CoV-2 in Norway, June 2022 – March 2023

**DOI:** 10.1101/2023.04.27.23289199

**Authors:** Ettore Amato, Susanne Hyllestad, Petter Heradstveit, Petter Langlete, Line Victoria Moen, Andreas Rohringer, João Pires, Jose Antonio Baz Lomba, Karoline Bragstad, Siri Laura Feruglio, Preben Aavitsland, Elisabeth Henie Madslien

## Abstract

**Background:** During the COVID-19 pandemic, wastewater-based surveillance gained great international interest as an additional tool to monitor SARS-CoV-2. In autumn 2021, the Norwegian Institute of Public Health decided to pilot a national wastewater surveillance (WS) system for SARS-CoV-2 and its variants between June 2022 and March 2023. We evaluated the system to assess if it met its objectives and its attribute-based performance.

**Methods:** We adapted the available guidelines for evaluation of surveillance systems. The evaluation was carried out as a descriptive analysis and consisted of the following three steps: (i) description of the WS system, (ii) identification of users and stakeholders, and (iii) analysis of the system’s attributes and performance including sensitivity, specificity, timeliness, usefulness, representativeness, simplicity, flexibility, stability, and communication. Cross-correlation analysis was performed to assess the system’s ability to provide early warning signal of new wave of infections.

**Results:** The pilot WS system was a national surveillance system using existing wastewater infrastructures from the largest Norwegian municipalities. We found that the system was sensitive, timely, useful, representative, simple, flexible, acceptable, and stable to follow the general trend of infection. Preliminary results indicate that the system could provide an early signal of changes in variant distribution. However, challenges may arise with: (i) specificity due to temporary fluctuations of RNA levels in wastewater, (ii) representativeness when downscaling, and (iii) flexibility and acceptability when upscaling the system due to limited resources and/or capacity.

**Conclusions:** Our results showed that the pilot WS system met most of its surveillance objectives. The system was able to provide an early warning signal of 1-2 weeks, and the system was useful to monitor infections at population level and complement routine surveillance when individual testing activity was low. However, temporary fluctuations of WS values need to be carefully interpreted. To improve quality and efficiency, we recommend to standardise and validate methods for assessing trends of new waves of infection and variants, evaluate the WS system using a longer operational period particularly for new variants, and conduct prevalence studies in the population to calibrate the system and improve data interpretation.

## Background

The concept of wastewater surveillance (WS) for infectious diseases is based on the evidence that some infectious agents are being excreted through urine and faeces from infected persons, including before start of symptoms and from asymptomatic cases. Therefore, WS could provide the first signal of change in disease trends as it is also at the lower level of the surveillance pyramid (1). During the COVID-19 pandemic, wastewater-based epidemiology gained great international interest as an additional tool to detect signals of SARS-CoV-2 transmission in communities and monitor trends in defined population areas to inform COVID-19 testing policies and mitigation measures (2, 3). WS has also been applied in the monitoring of SARS-CoV-2 trends and variants among travellers at international airports, passenger aircraft and cruise ships (4, 5, 6, 7). Moreover, several studies have demonstrated that community-wide molecular analysis of wastewater samples can be used to track SARS-CoV-2 variants and support the identification of potential new emerging variants (8, 9, 10). Following the European Commission’s recommendation to the Member States in spring 2021, several EU member countries have initiated, implemented, or established monitoring of SARS-CoV-2 and its variants in wastewater (11, 12).

In Norway, surveillance of COVID-19 has largely been based on registration of all individual test results in the Norwegian Surveillance System for Communicable Diseases (MSIS). However, a change in testing strategy (from PCR-based to self-testing using antigenic tests) in autumn 2021 resulted in a considerable proportion of test results not being registered in MSIS and raised the need to strengthen the national surveillance systems for SARS-CoV-2 (13). Consequently, the Norwegian Institute of Public Health (NIPH) decided to pilot a national WS system for SARS-CoV-2 to assess its usefulness and performance as a complementary tool to monitor SARS-CoV-2 and its variants.

This is the first time that WS has been tested by national health authorities in Norway as an operational monitoring system in connection with outbreaks or epidemics. Therefore, a thorough evaluation of the system’s quality and performance is relevant to identify learning points and assess potential for future use. Moreover, despite the increasing number of publications and reviews, there is still a lack of studies evaluating the usefulness and performance of WS systems (14).

Therefore, the aim of this study was to evaluate the pilot WS system for SARS-CoV-2 in Norway in order to assess the performance of the pilot WS in relation to the public health-specific objectives of the system and describe the advantages and challenges of WS compared to existing national surveillance systems.

## Methods

### Study design

We adapted the guidelines for evaluation of surveillance systems given by the European Centre for Disease Prevention and Control (ECDC) (15) and the U.S. Centers for Disease Prevention and Control (CDC)(16) to evaluate the performance of the pilot WS system in Norway. The evaluation was carried out as a descriptive analysis and consisted of the following three steps: (i) description of the WS system, (ii) identification of end-users and stakeholders and (iii) analysis of the system’s attributes and performance (sensitivity, specificity, timeliness, usefulness, representativeness, simplicity, flexibility, stability, and communication).

### Description of the wastewater surveillance system

We described the WS system in terms of pilot surveillance objectives, project timeline, WS structure and network, and participating wastewater sampling sites and municipalities.

### Identification of end-users and stakeholders

End-users and stakeholders were categorized in the following 3 groups: (i) wastewater treatment plants and operators at local level, (ii) public health authorities at local level, and (iii) public health authorities including risk assessors and managers at national level. In addition, the national authority responsible for wastewater legislation and environmental monitoring was consulted.

### Analysis of the system’s attributes and performance

The evaluation of the WS system focused on the attributes described below (Table 1). The attributes definitions were adapted from the ECDC and CDC guidelines (15, 16) to fit the purpose of the WS system. Our evaluation was based on descriptive comparison of results obtained from the WS with other relevant clinical indicators available during the pilot period (June 2022 – March 2023), and feedback from end-users, stakeholders and NIPH’s experts. Cross-correlation analysis was performed to assess the wastewater systems’ ability to provide early warning signal of new waves of infection. The analysis was performed using time series data. The time series included data from the main wave (from week 33, 2022 to week 10, 2023) which was divided into periods of increasing and decreasing trends, respectively.

**Table 1.**
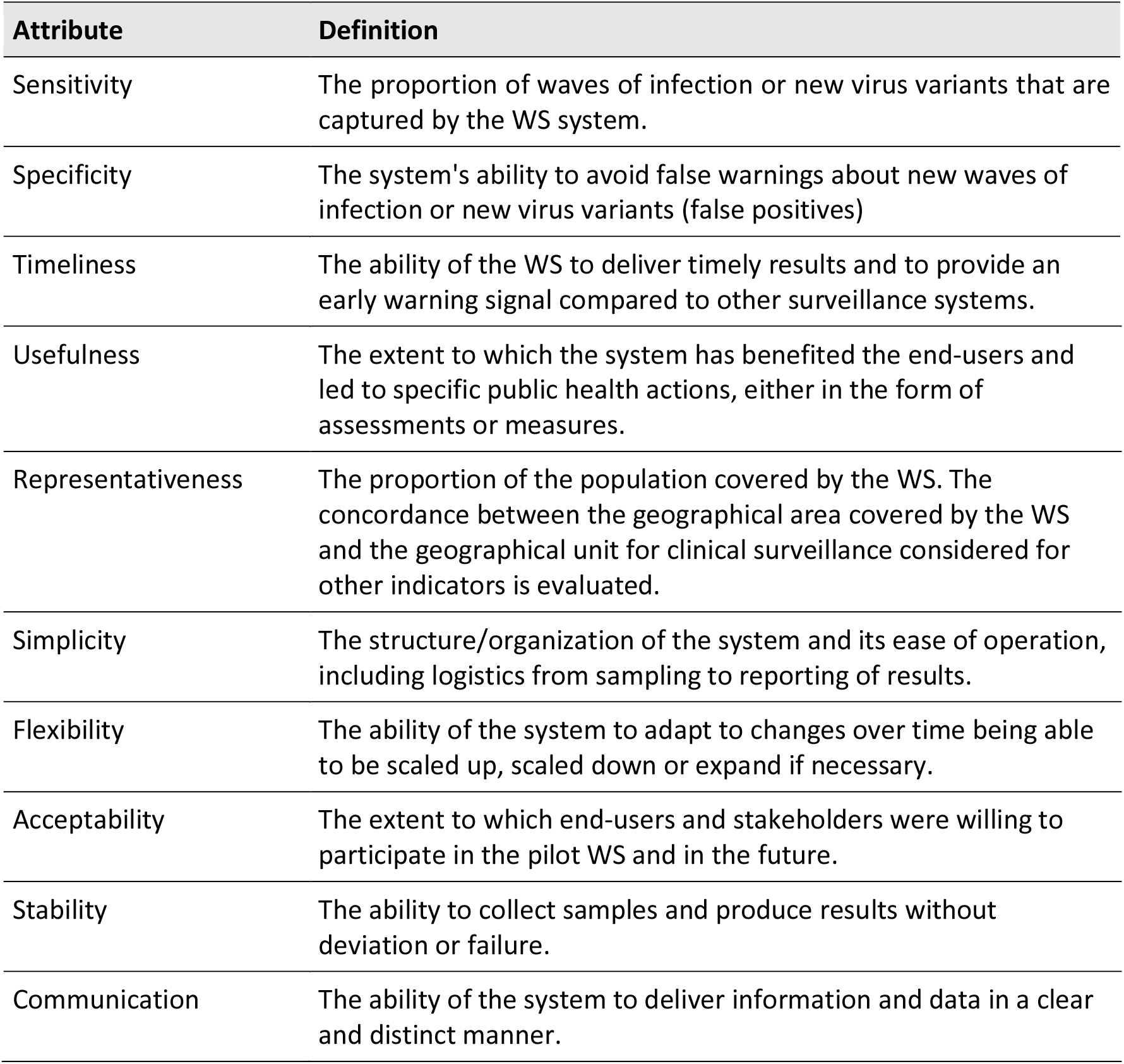
Wastewater surveillance attributes and their definitions as used in this study, adapted from ECDC and CDC guidelines

### Sources used for the surveillance evaluation

#### Survey collecting feedback from end-users and stakeholders

A questionnaire was developed and sent to involved stakeholders and participants of the pilot WS in February 2023 (Section A, Supplementary information). The questions were adapted to each end-user category and covered topics such as cooperation, communication, future areas of use and surveillance attributes. The information was collected, aggregated, and analysed based on feedback received. Experiences from NIPH’s project team and experts were also collected to describe the pilot wastewater system and its technical performance.

#### NIPH’s wastewater surveillance weekly reports

The weekly WS reports (17) included information and results produced by the WS system such as quantitative determination of SARS-CoV-2 using reverse transcription-quantitative polymerase chain reaction (RT-qPCR) and variant screening including results from both RT-PCR for specific variant mutations and from deep-sequencing analysis on parts of the Spike protein using Nanopore technology. These reports included data from the wastewater surveillance, the Emergency Preparedness Register for COVID-19 (Beredt C19) including MSIS and national registries on intensive care unit (ICU) and hospitalizations. The ratio between the concentration of genetic material (gene copies) for SARS-CoV-2 and the reference control pepper mild mottle virus (PMMoV) from wastewater samples was calculated. The results for each sample point were weighted according to the population size that belonged to the wastewater plant in an average at national level.

#### NIPH weekly reports on COVID-19, influenza, and other respiratory diseases

These reports included results reporting signals from other clinical indicators used by NIPH for the COVID-19 surveillance (13). The sources used for these reports included sequence data from the National Virological SARS-CoV-2 Surveillance Program and the clinical surveillance data form the Emergency Preparedness Register for COVID-19 (Beredt C19) which collects data from MSIS and the national registries on ICU and hospitalizations.

## Results

### Description of the pilot wastewater surveillance system

The pilot WS system is a national surveillance system, which aims to provide information on the occurrence and circulation of SARS-CoV-2 and its variants in a defined population.

#### Surveillance objectives

The public health-specific objectives of the WS system for SARS-CoV-2 in Norway were to (i) describe trends of virus circulation, including its variants in the population over time and space, (ii) provide an early detection of change in infection trends in the population compared to other national COVID-19 surveillance systems and indicators, and (iii) detect and monitor emerging variants of public health relevance.

#### Pilot Wastewater Surveillance timeline

The pilot WS system was operational from 1^st^ June 2022, with a tentative trial period of 6 months. In December 2022, the pilot surveillance was extended in a scaled-down version until March 2023 to gather additional experience covering the winter season and data useful to perform an evidence-based evaluation of the surveillance and its performance.

#### Wastewater Surveillance structure and network

The system used existing municipal wastewater infrastructures. A network of reference contact persons was established for (i) the managers of enrolled wastewater treatment plants, (ii) the municipal doctors in participating municipalities and iii) an outsourced private laboratory (contract laboratory) for the RT-qPCR analysis of wastewater samples. The contract laboratory was responsible for the logistics and initial analysis of the wastewater samples, including quantitative detection of SARS-CoV-2 RNA, quantitative detection of Pepper Mild Mottle Virus RNA (PMMoV) and PCR screening for a pre-defined set of mutations frequently observed in known Variants of Concern (VOCs). Frozen samples containing extracted nucleic acids were then shipped to the national reference laboratory at NIPH for further variant analysis and deep-sequencing of the Spike protein. The results from the contract laboratory were shared through an online platform and then processed by NIPH before being compared to other surveillance systems data and indicators. The municipal doctors in the participating municipalities (Oslo, Bergen, Trondheim, Tromsø, and Oslo airport area) were involved in the coordination, sharing of results and communication at the local level. NIPH was responsible for the overall administration and coordination of the project, analysis and interpretation of data and communication and reporting at national and European level.

#### Sampling procedure and sites included in the pilot wastewater surveillance

From June to November 2022, wastewater treatment plants in the largest urban municipalities representing each region participated in the pilot WS. These include wastewater treatment plants placed in Oslo (n=2), Bergen (n=4), Trondheim (n=2), and Tromsø (n=3). In addition to the largest urban municipalities representing for each Norwegian region, the Oslo airport area (the airport with the highest international influx in Norway) was included to detect new variants of public health relevance. From December 2022 to March 2023, the pilot surveillance was downscaled from 12 to 5 wastewater treatment plants placed in Oslo (n=2), Bergen (n=1), Trondheim (n=1) municipalities and Oslo airport area (n=1). Sampling was carried out one to two times per week by the wastewater treatment plants’ personnel. All samples were collected from untreated water at the inlet of the plant. Most samples (68%) were collected using 24-72h flow-proportional composite samples, 19% were collected using time-proportional composite samples and 13% were collected by grab sampling. Sample material was transferred to 1L bottles and shipped cooled on ice to the contract laboratory.

### Identification of end-users and stakeholders

The list of identified end-users and stakeholders for each category is presented in Table 2.

**Table 2.** Identified end-user and stakeholders for each category of the pilot wastewater surveillance.

| Category | Identified end-users and stakeholders |
| --- | --- |
| Wastewater treatment plants | Contact persons from participating wastewater treatment plants |
| Local public health authorities | Municipal doctors from the 5 participating municipalities |
| National public health authorities and experts on infection control and preparedness | The Directorate of Health and the Norwegian Institute of Public Health |
| National authority responsible for wastewater legislation and environmental monitoring | The Norwegian Environment Agency |

### Analysis of the surveillance system’s attributes and performance

#### Sensitivity - waves of infection

During the pilot period (June 2022 - March 2023), we observed two waves of infection in Norway and the beginning of a third wave (Figure 1). The first wave from week 22 to week 38 (2022) and the second wave was recorded from approximately week 40 (2022) to week 4 (2023), while the beginning of a third wave started around week 5 (2023). Comparing with clinical indicators (such as registered COVID-19 cases, hospitalizations, and ICU admissions), all waves were captured by the WS system, indicating that the system had a similar sensitivity compared to other surveillance systems both at national and local level (Figure 1 and 2).

**Figure 1.**
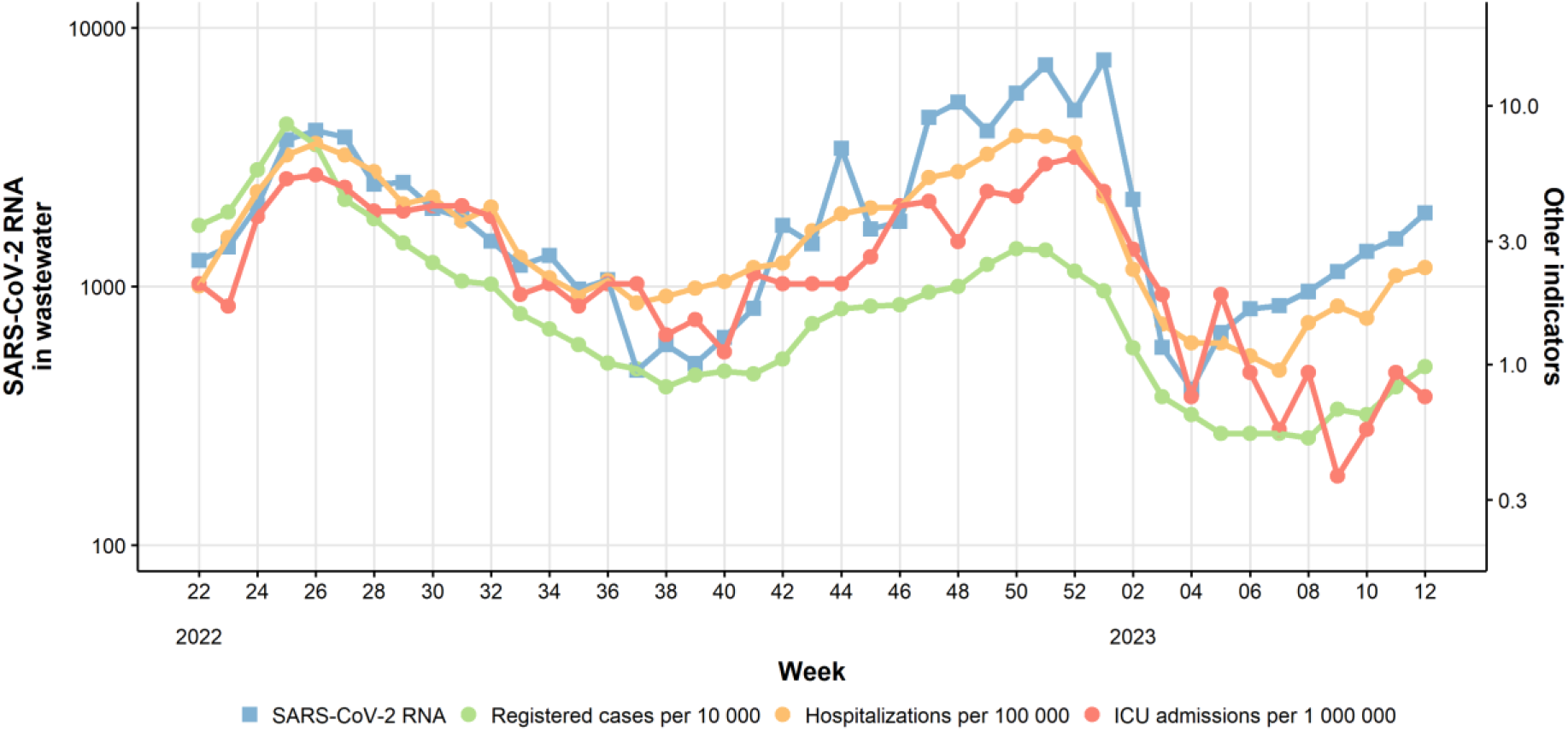
Weekly level of SARS-CoV-2 RNA detected in wastewater in Norway (blue line), compared with clinical indicators for COVID-19. SARS-CoV-2 RNA levels in wastewater are population-weighted and PMMoV-normalized. Note: the wastewater data are based on results of samples taken at selected locations, while the clinical indicators are based on data from national registries (Beredt-C19).

**Figure 2.**
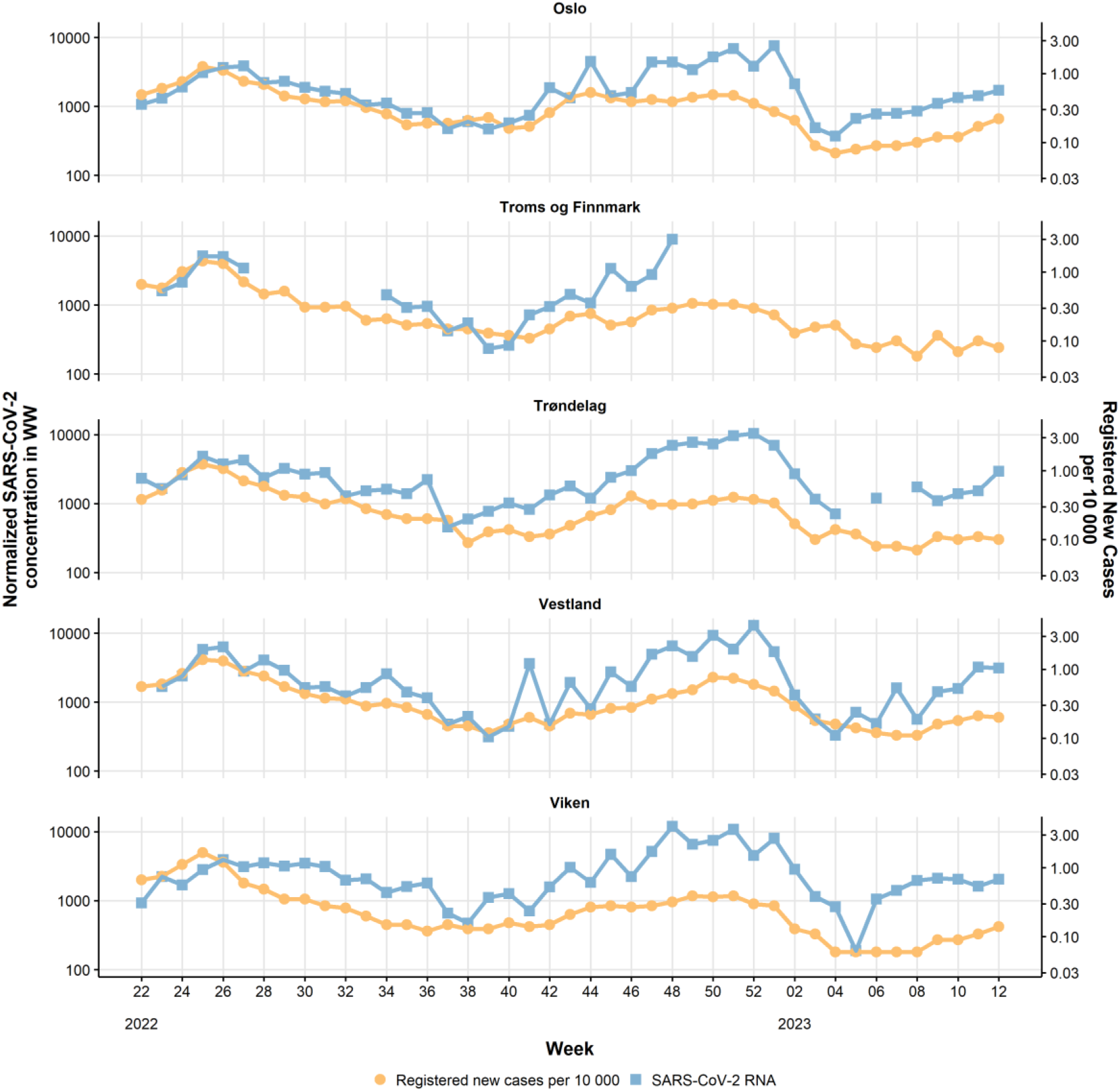
Weekly concentration of SARS-CoV-2 RNA in wastewater (blue) compared to weekly registered cases expressed as incidence per 10,000 inhabitants (orange). SARS-CoV-2 RNA levels in wastewater are population-weighted and PMMoV-normalized. Note: the wastewater data are based on results of samples taken at selected locations, while the data on registered cases are based on national registries (Beredt C19).

#### Specificity - waves of infection

Compared to the Beredt C19 clinical indicators, we observed that the WS system gave signals of temporary fluctuations over the weeks that were not otherwise captured. These fluctuations made the interpretation of the wastewater signals challenging, since it was unclear in a real-time situation whether a weekly increased value represented a real increase or was a consequence of random fluctuations, or measurement errors which could occur at different stages of the process from sampling to final result.

#### Timeliness - waves of infection

The results provided by the WS system correlated with clinical indicators related to this wave but did not give an early signal of downward trend compared to registered clinical cases. During the autumn of 2022, we observed the beginning of a new wave of infection (Figure 1). In the NIPH’s COVID-19 weekly report for weeks 37 and 38 (18, 19), a “slightly decreasing or flat trend” was generally reported, while the WS system reported a slightly increasing trend. In the following weeks 39 and 40, the COVID-19 report (20, 21) was on “stable” spread of infection, while the WS system reported a “tendency to increase”. The WS system gave an earlier warning of the new wave of infection than clinical indicators. During increasing trends, we found the highest correlation between the wastewater data and the clinical indicators at a lead time of 1-2 weeks (Figure 1S, Section B, Supplementary material). During the decreasing trends the correlation was highest at a lead time around zero (Figure 2S, Section B, Supplementary material).

Results were published through weekly reports using different communication channels. Overall, completeness of weekly results from the WS system was 1-7 days earlier compared to the clinical registry-based surveillance systems. In Figure 3, we show an example of surveillance results from different systems updated routinely during a week, particularly how the results would be updated on Mondays (A), Wednesdays (B) and Fridays (C).

**Figure 3.**
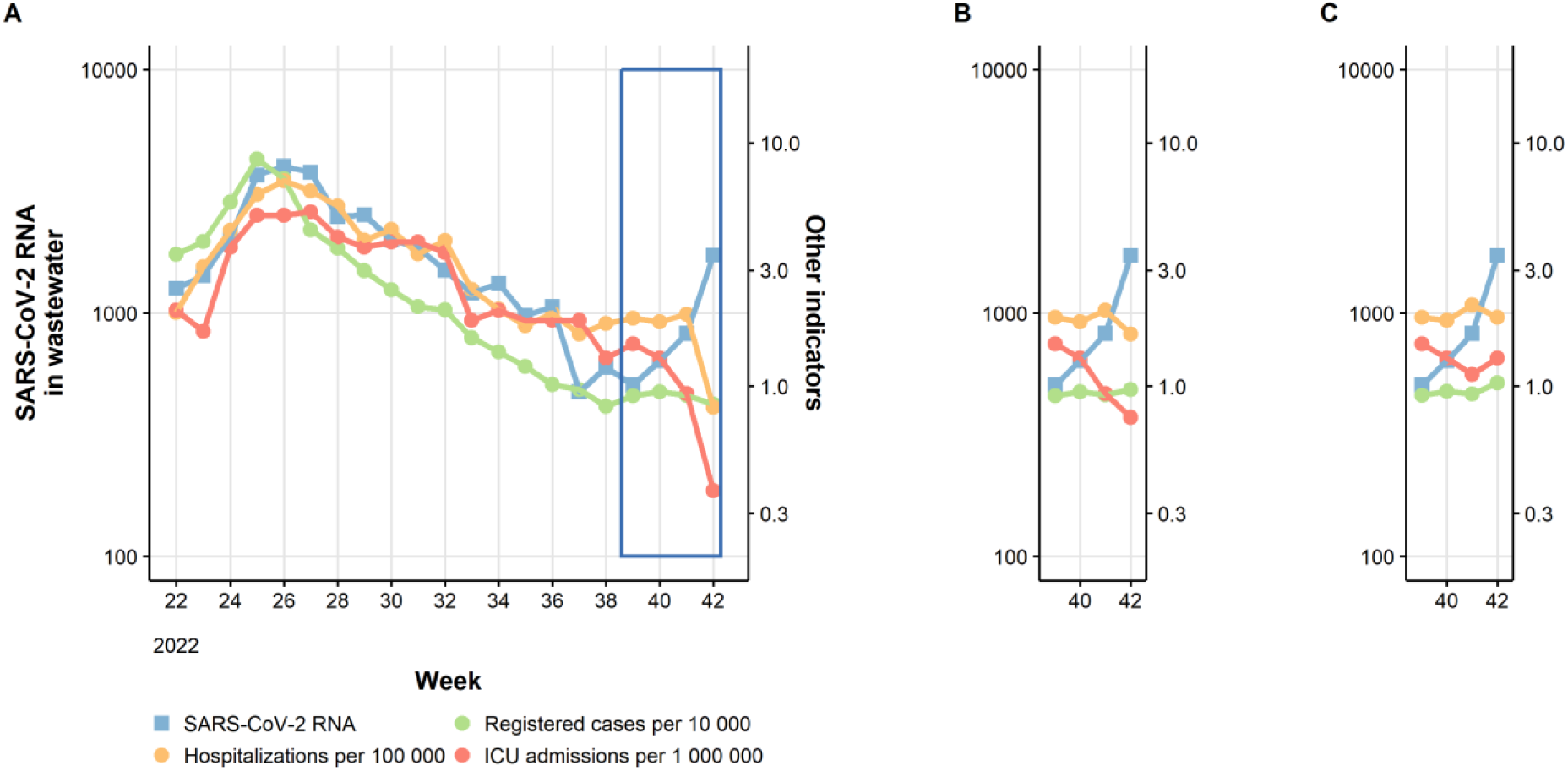
Timeliness of gathering complete results by different surveillance systems over a defined week: A) WS data updated on Mondays (blue), B) hospitalizations (yellow) and registered cases (green) data updated on Wednesdays and C) ICU admissions (red) data updated on Fridays.

#### Sensitivity, specificity and timeliness of wastewater surveillance for variants detection

While mutational PCR screening was performed by the contract laboratory and reported for the entire study period, reporting of sequencing results started only at a late stage (week 47). Overall, mutational PCR screening results showed concordance with the signals generated from the clinical variants surveillance (13, 17). However, the PCR screening method did not have sufficient discriminatory power to detect and identify specific variants over time. Preliminary sequencing data, both real-time and retrospective data (Supplementary Figures S3), suggest that signals of new key-mutations and changes in variant distribution from the WS system preceded signals from clinical variant surveillance approximately by 1-2 weeks.

However, additional data would be needed to conduct a proper evaluation of the WS systems’ real-time performance in terms of sensitivity, specificity, and timeliness for new variants of public health relevance.

### Usefulness

The overall opinion reported by end-users through the evaluation survey was that the WS system was useful to follow the general trend of infections, both at national and local level. Surveys results showed that all end-users at municipal level would have benefited from the WS system if it had started earlier in the pandemic, e.g., to monitor the rate of transmission, especially in periods with extensive use of self-tests, or to inform potential easing or strengthening of measures at population level.

Three out of five municipalities reported that the results were used to assess the epidemiological situation or need for infection control measures, one out of five indicated that the results could have been more useful if the system had started earlier during the pandemic and the last believed that it could be relevant to use this system in an assessment of measures if the situation had changed in a negative direction. The overall feedback from national authorities and stakeholders was that the results of the pilot WS were used as one of several indicators to assess the infection situation nationally. It was considered particular useful in a phase of the pandemic where testing activity, and hence the reliability of traditional surveillance systems, has been significantly lower than earlier in the pandemic. Several end-users addressed the importance of being able to monitor other pathogens to increase the future usefulness of the system. Signals from the WS have varied from week to week, which increases the risk of misjudgement of the trends. For the future, it will be relevant to identify sources of these variations and how to minimize them, as well as establish standardized guidelines for assessing and communicating trends and uncertainties.

### Representativeness

The selected 12 wastewater treatment plants included in the pilot’s first phase (June-November 2022) covered approximately 30% of the Norwegian population. The scaled down phase of the pilot (December 2022-March 2023) included only five wastewater treatment plants with a coverage of around 25% of the population. We have simulated a further scale down of the pilot including only Oslo municipality and the airport area with a coverage of around 22%. The trend analysis considering these three scale down situations showed that the national trend is similar in all scenarios, but the results were less reliable at the local level.

### Simplicity

The WS system required the involvement of external actors. For this reason, coordination was required to establish collaborations and run the system together with actors not usually involved in traditional clinical surveillance systems. Considering the scale of the pilot system, the operations were manageable both in terms of coordination, resource allocation and communication between stakeholders and end-user. Handling of data and processing of results was largely automated. The complexity could increase in an upscaling scenario due to the increased number of wastewater treatment plants or municipalities involved.

### Flexibility

The system was flexible to the extent that the number of sampling locations and sampling frequency can be scaled up and down when needed. During the operational period of the pilot, we were able to test the ability to scale down, which proceeded without major challenges. Scaling up requires that the treatment plants and the laboratory have sufficient capacity. Feedback from operators of the treatment plants suggested that capacity-related challenges may arise for several of them if there is a need for upscaling. Shortages of personnel, logistics and time pressure were mentioned as the biggest possible challenge. Different end-users expressed interest in using the WS system to monitor other pathogens in addition to SARS-CoV-2 in the future

### Acceptability

Both local health authorities and the wastewater treatment plants’ operators largely expressed a willingness to continue contributing to the WS system, taking into account their available resources and capacity. One of the municipalities participating in the pilot did not have the capacity to participate further.

### Stability

The system showed stability in terms of delivering regular results for assessment of national trends. However, we occasionally experienced deviations. Deviations could occur for several reasons and at different stages in the WS system. The most important reasons we have registered were: (i) sampling, such as lack of capacity to take samples, which sometimes resulted in one sample per week instead of two. Capacity challenges were often linked to holiday closures and public holidays, (ii) logistics such as delays and deviations in connection with the collection and transport of samples. (iii) analysis deviations such as inhibition of the PCR analysis.

### Communication

All end-users and stakeholders reported that the results were clearly presented and the content sufficient for their needs. As additional feedback, end-users suggested adding more information on virus variants and proposed to include an indicator of the burden on the primary healthcare service, together with the hospital’s admission figures. End-users of the system at municipal level suggested that direct reporting to the municipal contact person was the preferred channel for communicating and accessing results rather than visiting the NIPH’s website to check the published reports. The frequency of reporting, once per week, was considered adequate during the study period.

## Discussion

Our study provides a detailed evaluation of the pilot WS system for SARS-CoV-2 including its performance in Norway during the study period (June2022 - March 2023). The Norwegian SARS-CoV-2 WS system was established at a late stage of the pandemic where individual clinical testing activity captured by COVID-19 national surveillance systems was decreasing due to a change of national testing strategy. Thus, there was a need to implement new systems to strengthen the national surveillance of SARS-CoV-2.

When a new surveillance system is implemented or established, periodic evaluation of such system is useful to ensure that the system fulfils its public health surveillance objectives and to identify areas of improvement (16). Guidelines for the evaluation of surveillance systems are available but largely tailored for evaluating clinical data and indicators using an attribute-oriented approach (15).

Evaluation guidelines adapted to wastewater-based public health surveillance systems are currently lacking. In this study, we share our experience in adapting the available ECDC and CDC guidelines to a pilot wastewater-based surveillance system of SARS-CoV-2 in Norway and assess the performance of this system during the study period.

Our assessment on the sensitivity, specificity and timeliness attributes was performed through descriptive analysis of WS data and comparison with relevant clinical indicators that were used for COVID-19 surveillance purposes during the pilot period in Norway. The definition on sensitivity and specificity were adapted for event-based surveillance, where we considered a new wave of infection or the introduction of a new virus variant as the “event” (15). While for timeliness we specifically evaluated the ability of the system to deliver timely and complete results and to provide an early warning signal to reflect the speed between steps in public health surveillance (15). Since the pilot was performed during the late phase of the pandemic where no major infection control measures were applied to the Norwegian population at national or local level, it was not possible to evaluate the reactivity of the system which reflects the delay before public health actions were initiated (15). During the autumn 2022, Norway experienced a new wave of infection. In this case, the WS system gave an approximately 1-2 weeks early warning signal for the new wave of infection compared with NIPH’s clinical surveillance indicators. This is also in line with the early warning window reported in literature (14). However, we did not find that the WS system could detect an early signal on steady state or declining trends, which could have been useful to forecast decrease in COVID-19 related illness, medical consultations, and hospitalizations (22). Although the WS system was able to capture national waves of infection and trends similarly to other indicators, some fluctuations were observed during the study period. Therefore, it is important to interpret weekly results carefully and further investigate the factors influencing these deviations to reduce uncertainties, such as population dynamics, in-network characteristics, sampling strategy and sample analysis (23). For example, simulations from a recent Danish study reported that a large variation in the viral concentration per gram of faeces between infected individuals results in a large variability in the concentrations found in wastewater, especially when the number of shedders is low (24). Increasing sampling frequency would presumably lower the impact of random fluctuations in RNA levels, however this would demand more resources.

Regarding PCR-screening of signature mutations of VOCs, the analysis of pooled samples from each municipality carried out by the contract laboratory showed concordance with variant results from the National Virological SARS-CoV-2 Surveillance Program. This analysis gives a preliminary indication of whether certain known signature mutations are present or absent in the population. These results could be useful to provide an early signal of changes in known variants given that the selection of mutations that are targeted are relevant and can be continuously and timely adapted according to the evolving needs. However, the method has a very low discriminatory power in terms of providing information on which variants are present and their relative distribution, and the results should therefore be carefully interpreted and complemented by sequencing analysis, especially to detect also unknown variants. Thus, as a stand-alone method, PCR-based mutational screening of wastewater samples is not suitable for determining the true variant prevalence or assessing variant shifts over time.

Sequencing results of pooled samples from each municipality became available after week 47 (2022) and relative prevalence of variants after week 5 (2023) due to extensive work in developing and establishing suitable methods for analysing SARS-CoV-2 variant distribution in wastewater samples. Preliminary sequencing results suggest that signals on new mutations and variants were detected approximatively 1-2 weeks earlier than observed through the National Virological SARS-CoV-2 Surveillance Program. However, additional data would be needed to perform a thorough evaluation of the system’s performance in terms of its variant surveillance objectives. Variant classification (e.g. using the Pango system) (25) from wastewater samples was proven to be difficult, especially when only including part of the Spike protein for the sequencing analysis. The development and validation of sequencing methodology and robust bioinformatic pipelines suitable for complex wastewater samples could be resource-intense and needs to be considered when planning or establishing the WS system, particularly to be able to detect unknown future mutations.

All end-users at municipal level reported that the system has been useful during the pilot phase, and they would have also benefited from it if it had started earlier in the pandemic. However, while objective thresholds for clinical indicators were used to implement infection control measures both at national and municipal level during the earlier phase of the pandemic (e.g., COVID-19 incidence of positive cases at national and municipal level or hospitalization and intensive care unit occupancy rate) (26), there is still a need for guidelines on how to identify control measures’ thresholds for WS. Therefore, interpreting surveillance data and integrating different data sources considering each system’s limitation is essential from the public perspective to provide accurate advice and implement proportionate control measures, particularly when those have a high social and economic impact on the population.

Regarding representativeness, we evaluated how the WS system described the occurrence of a health-related event over time and its distribution in the population by place. An evaluation of the distribution by person was not possible due to the pooled nature of wastewater samples. We also evaluated the concordance of results from different systems considering the geographical area covered by the WS and the one used for other clinical indicators. The WS pilot study aimed at covering the highest percentage of the Norwegian population representative for each region considering also available resources within a framework of the surveillance needs at the national level. Our results showed that the disease trends over time were similar in different down-scaled scenarios, however the system lost its regional geographical representativeness which was not in line with the surveillance objectives.

Moreover, the catchment area of wastewater treatment plants rarely corresponds to the municipal or county boundaries. These aspects need to be considered when implementing WS for epidemiological purposes to integrate and compare results with clinical indicators, particularly at local level. Although, it is worth noting that results from WS are independent of individual testing or reporting of symptoms. Therefore, the WS system can be considered an unbiased source of information compared to register-based clinical surveillance to follow-up the infection transmission at population level over time, particularly in case of low individual testing or change of national testing strategies. The interpretation of results from the WS system would probably have been improved if we had been able to compare them to results from a study of infection prevalence in the same geographical areas. Unfortunately, no such studies were performed in Norway.

Although the system proved to be simple, flexible, acceptable, and stable by end-users and stakeholders during the pilot phase, some resource-related challenges might arise in a possible upscale scenario. These challenges are relevant when establishing or implementing WS systems, particularly in high demand situations (e.g., during the acute phase of a pandemic).

Our results also highlighted the importance of timely and direct communication with end-users and stakeholders involved in the pilot WS system, as well as the usefulness of interpreting the epidemiological situation using multiple data sources.

### Regulatory framework of wastewater surveillance for public health purposes

Sampling of wastewater for monitoring diseases in the population is not directly regulated in the Norwegian legislation. In October 2022, changes were proposed to the EU Directive for “Urban wastewater treatment” with a new section (Article 17-Urban wastewater surveillance) which deals with the use of wastewater for public health purposes (27). The proposal, if it is adopted, entails, among other things, that the EU/EEA countries must have established a coordinating structure (between environmental and health authorities) for monitoring public health parameters in wastewater (covering 70% of the population) by 1 January 2025. The Norwegian Environment Agency is responsible for following up the proposed wastewater directive in Norway.

### Outlook

This evaluation has identified the need for an update of the international guidelines used for the evaluation of surveillance systems for public health purposes which should include both clinical and environmental indicators. Moreover, standardized analytical methods used for WS and a harmonized approach for surveillance evaluations would be useful to compare the results, performance and added value of WS in different countries. This aspect is considered particularly relevant when results are shared through international platforms and dashboards. In addition, further studies exploring the sustainability and use of WS for other emerging or relevant public health threats can add to the discussion on the usefulness of WS for public health purposes, particularly when the surveillance is focusing on aspects related to the interphase between animal, human and environmental health, using the ‘One Health’ approach.

### Limitations of the surveillance evaluation

Although this study can be a valuable example when planning an evaluation for WS system for public health purposes, we have identified several limitations that must be addressed to improve the quality of such evaluations in the future. First, a more thorough statistical analysis was not possible due to the lack of a ‘gold standard’ indicator on the incidence of the disease in the population during the pilot period. Secondly, the pilot started at a time when a new wave of infections was already in the starting phase (week 22/2022). Therefore, it was not possible to assess if the system would have warned of this first wave earlier than the Beredt C19 indicators. Third, although additional questions in the survey could have increased the knowledge of the WS system’s performance, we have limited those questions to ensure a good response rate from end-users and stakeholders. In addition, we did not perform a cost-benefit analysis since this would require a different study design and evaluation method which was outside our scope. It was also not possible to evaluate the laboratory testing capacity in case of an upscaling scenario, since we have outsourced the laboratory responsible for testing the wastewater samples under specific contract requirements. Moreover, the variant’s sequencing analysis was operational only during the last weeks of the pilot, therefore it was not possible to thoroughly evaluate the sensitivity, specificity and timeliness for the surveillance of variants. Finally, the evaluation is limited to the pilot settings and does not include experiences from local surveillance initiated by individual small municipalities. We have also evaluated the performance of the WS system based on its routine sampling and analysis procedures, however adjustment of these factors could increase or decrease the performance of WS system and surveillance’ attributes.

## Conclusions and recommendations

As result of this surveillance evaluation, we observed that: (i) the WS system met most of its surveillance objectives, (ii) the WS system provided an early warning signal of 1-2 weeks for a new wave of infection compared to other clinical indicators, (iii) temporary fluctuations of wastewater values can cause noisy signals that makes the interpretation of trends challenging and increasing the risk of false alert, iv) preliminary results indicate that the WS system sequencing of wastewater samples could provide an early signal of selected SARS-CoV-2 key mutations and changes in variant distribution over time compared to the clinical SARS-CoV-2 variants surveillance system (National Virological SARS-CoV-2 Surveillance Program), but additional data would be needed to conduct a proper evaluation of the WS systems’ real-time performance for variants’ detection and (v) the system is seen as a useful tool by local and national health authorities to monitor the infections at population level when individual testing activity is low and complement other surveillance systems. Based on these results, the WS system has been extended beyond its pilot phase in Norway and its future use will be evaluated based on continuous assessments of national public health needs and available resources.

To improve the quality and efficiency of the WS system, we would recommend to standardize and validate methods for assessing trends of new waves or virus variants, evaluate the WS system for sensitivity, specificity and timeliness for variants using a longer surveillance operational period, identify the causes of trend fluctuation to minimize the challenges in the interpretation of WS signals, and conduct prevalence studies in the population to calibrate WS data and improve the interpretation of data.

## Supporting information

Supplementary material_Evaluation Wastewater surveillance_Norway

## Data Availability

The dataset analysed in the study contains individual-level data from various central health registries or laboratory databases. Only fully anonymized data (i.e., data that are neither directly nor potentially indirectly identifiable) are permitted to be shared publicly. Therefore, legal restrictions prevent the researchers from publicly sharing the dataset used in the study. However, external researchers are freely able to request access to data as per normal procedure for conducting health research in line with national and European regulations concerning data protection.

## Abbreviations

WS: Wastewater surveillance
MSIS: The Norwegian surveillance system for communicable diseases
NIPH: Norwegian Institute of Public Health
ECDC: European Centre for Disease Prevention and Control
CDC: U.S. Centers for Disease Control and Prevention
PCR: Polymerase Chain Reaction
RT-PCR: Reverse Transcription Polymerase Chain Reaction
RT-qPCR: Quantitative Reverse Transcription Polymerase Chain Reaction
Beredt C19: Emergency Preparedness Register for COVID-19
ICU: Intensive Care Unit
PMMoV: Pepper Mild Mottle Virus
VOC: Variant of Concern
EU: European Union
EEA: European Economic Area

## Acknowledgements

We thank the municipal doctors and personnel at the wastewater treatment plants for their willingness to participate in the Norwegian wastewater surveillance pilot and for their valuable contributions and feedback during the evaluation study. We also thank Rasmus Kopperud Riis and Ignacio Garcia Llorente (NIPH) for their technical skills in developing a unique pipeline for wastewater genomics surveillance useful to produce virus variants data from wastewater samples, and Astrid Louise Løvlie (NIPH) and Aftab Jasir (EPIET/EUPHEM ECDC Fellowship Programme) for reviewing the manuscript. We also acknowledge Lasse Dam Rasmussen and Kristina Træholt Franck at Statens Serum Institut (SSI) in Denmark for valuable guidance during the start-up of the pilot, and Matthew Wade at the UK Health Security Agency for discussions during the planning phase of the WS evaluation.

## Author’s contributions

EA, SH, and EM designed the study. PH helped to develop the survey and correspondence with end-users and stakeholders. PL, LVM, AR, KB analysed and helped in interpreting the data from the surveillance systems. JP and JABL helped with technical inputs from the scientific literature. JP and PL prepared and harmonized the figures. SLF and PA helped in interpreting the surveillance evaluation results under the preparedness and public health perspective. EA and EM drafted the manuscript. All co-authors revised the manuscript and have approved the final version.

## Ethics approval and consent to participate

Approval was obtained from the Regional Committee for medical and healthcare research ethics (REK 454077) based on the Norwegian Health Research Act.

## Consent for publication

Not applicable.

## References

1. World Health Organization. Environmental surveillance for SARS-CoV-2 to complement public health surveillance - Interim Guidance. https://www.who.int/publications/i/item/WHO-HEP-ECH-WSH-2022.1. 2022. Accessed 27 Apr 2023.

2. Medema G, Been F, Heijnen L, Petterson S. Implementation of environmental surveillance for SARS-CoV-2 virus to support public health decisions: Opportunities and challenges. Curr Opin Environ Sci Health. 2020;17:49.

3. Centers for Disease Control and Prevention. National Wastewater Surveillance System (NWSS). https://www.cdc.gov/nwss/wastewater-surveillance.html. 2021. Accessed 27 Apr 2023.

4. Ahmed W, Bertsch PM, Angel N, Bibby K, Bivins A, Dierens L, et al. Detection of SARS-CoV-2 RNA in commercial passenger aircraft and cruise ship wastewater: a surveillance tool for assessing the presence of COVID-19 infected travellers. J Travel Med. 2020;27(5).

5. Medema G HL, Elsinga G, Italiaander R, Brouwer A. Presence of SARS-Coronavirus-2 RNA in Sewage and Correlation with Reported COVID-19 Prevalence in the Early Stage of the Epidemic in The Netherlands. Environ Sci Technol Lett. 2020.

6. Farkas K, Williams R, Alex-Sanders N, Grimsley JMS, Pântea I, Wade MJ, et al. Wastewater-based monitoring of SARS-CoV-2 at UK airports and its potential role in international public health surveillance. PLOS Glob Public Health. 2023;3(1):e0001346.

7. Agrawal S, Orschler L, Tavazzi S, Greither R, Gawlik BM, Lackner S. Genome Sequencing of Wastewater Confirms the Arrival of the SARS-CoV-2 Omicron Variant at Frankfurt Airport but Limited Spread in the City of Frankfurt, Germany, in November 2021. Microbiol Resour Announc. 2022;11(2):e0122921.

8. Kirby AE, Welsh RM, Marsh ZA, Yu AT, Vugia DJ, Boehm AB, et al. Notes from the Field: Early Evidence of the SARS-CoV-2 B.1.1.529 (Omicron) Variant in Community Wastewater - United States, November-December 2021. MMWR Morb Mortal Wkly Rep [Internet]. 2022 2022/01//; 71(3):103.

9. Izquierdo-Lara R, Elsinga G, Heijnen L, Munnink BBO, Schapendonk CME, Nieuwenhuijse D, et al. Monitoring SARS-CoV-2 Circulation and Diversity through Community Wastewater Sequencing, the Netherlands and Belgium. Emerg Infect Dis. 2021;27(5):1405.

10. Tiwari A, Adhikari S, Zhang S, Solomon TB, Lipponen A, Islam MA, et al. Tracing COVID-19 Trails in Wastewater: A Systematic Review of SARS-CoV-2 Surveillance with Viral Variants. Water [Internet]. 2023; 15(6).

11. European Commission. Commission recommendation of 17.3.2021 on a common approach to establish a systematic surveillance of SARS-CoV-2 and its variants in wastewaters in the EU. https://eur-lex.europa.eu/legal-content/EN/TXT/PDF/?uri=CELEX:32021H0472. 2021. Accessed 27 Apr 2023.

12. European Commission. EU Sewage Sentinel System for SARS-CoV-2, Digital European Exchange Platform (EU4S-DEEP). https://wastewater-observatory.jrc.ec.europa.eu/#/dashboards/2. Accessed 27 Apr 2023.

13. Norwegian Institute of Public Health. Weekly reports on COVID-19, Influenza and other respiratory infections. https://www.fhi.no/publ/2020/koronavirus-ukerapporter/. Accessed 27 Apr 2023.

14. Hyllestad S, Myrmel M, Lomba JAB, Jordhøy F, Schipper SK, Amato E. Effectiveness of environmental surveillance of SARS-CoV-2 as an early warning system during the first year of the COVID-19 pandemic: a systematic review. J Water Health. 2022;20(8):1223.

15. European Centre for Disease Prevention and Control. Data quality monitoring and surveillance system evaluation - A handbook of methods and applications. https://www.ecdc.europa.eu/en/publications-data/data-quality-monitoring-and-surveillance-system-evaluation-handbook-methods-and. 2014. Accessed 27 Apr 2023.

16. German RR, Lee LM, Horan JM, Milstein RL, Pertowski CA, Waller MN. Updated guidelines for evaluating public health surveillance systems: recommendations from the Guidelines Working Group. MMWR Recomm Rep. 2001;50(Rr-13):1.

17. Norwegian Institute of Public Health. Results from wastewater surveillance. https://www.fhi.no/en/hn/statistics/overvaking-smittsomme-sykdommer-i-avlopsvann/results-from-wastewater-surveillance/. Accessed 27 Apr 2023.

18. Norwegian Institute of Public Health. Results from SARS-CoV-2 wastewater surveillance. Week 37. https://www.fhi.no/contentassets/de5043048b0445f98feef0f7e215ff83/vedlegg/2022---uke-22-37-resultater-avlopsovervakingen.pdf. Accessed 27 Apr 2023.

19. Norwegian Institute of Public Health. Results from SARS-CoV-2 wastewater surveillance. Week 38. https://www.fhi.no/contentassets/de5043048b0445f98feef0f7e215ff83/vedlegg/2022---uke-22-38-resultater-avlopsovervakingen.pdf. Accessed 27 Apr 2023.

20. Norwegian Institute of Public Health. Results from SARS-CoV-2 wastewater surveillance. Week 39. https://www.fhi.no/contentassets/de5043048b0445f98feef0f7e215ff83/vedlegg/2022---uke-22-39-resultater-avlopsovervakingen.pdf. Accessed 27 Apr 2023.

21. Norwegian Institute of Public Health. Results from SARS-CoV-2 wastewater surveillance. Week 40. https://www.fhi.no/contentassets/de5043048b0445f98feef0f7e215ff83/vedlegg/2022---uke-22-40-resultater-avlopsovervakingen.pdf. Accessed 27 Apr 2023.

22. Schenk H, Heidinger P, Insam H, Kreuzinger N, Markt R, Nägele F, et al. Prediction of hospitalisations based on wastewater-based SARS-CoV-2 epidemiology. Sci Total Environ. 2023;873:162149.

23. Wade MJ, Lo Jacomo A, Armenise E, Brown MR, Bunce JT, Cameron GJ, et al. Understanding and managing uncertainty and variability for wastewater monitoring beyond the pandemic: Lessons learned from the United Kingdom national COVID-19 surveillance programmes. J Hazard Mater. 2022;424(Pt B):127456.

24. Nauta M, McManus O, Træholt Franck K, Lindberg Marving E, Dam Rasmussen L, Raith Richter S, et al. Early detection of local SARS-CoV-2 outbreaks by wastewater surveillance: a feasibility study. Epidemiol Infect. 2023;151:e28.

25. Centers for Disease Control and Prevention. SARS-CoV-2 Variant Classifications and Definitions. https://www.cdc.gov/coronavirus/2019-ncov/variants/variant-classifications.html. Accessed 27 Apr 2023.

26. Norwegian Institute of Public Health. Handbook for detection, assessment and handling of COVID-19 outbreaks in the municipality. Chapter 4. Risk assessment. https://www.fhi.no/nettpub/overvaking-vurdering-og-handtering-av-covid-19-epidemien-i-kommunen/ti-trinn2/4-risikovurdering/. Accessed 27 Apr 2023.

27. European Commission. Proposal for a DIRECTIVE OF THE EUROPEAN PARLIAMENT AND OF THE COUNCIL concerning urban wastewater treatment. https://eur-lex.europa.eu/legal-content/EN/TXT/HTML/?uri=CELEX:52022PC0541. Accessed 27 Apr 2023.

