## Supplementary material_Evaluation Wastewater surveillance_Norway for "Evaluation of the Pilot Wastewater Surveillance for SARS-CoV-2 in Norway, June 2022 – March 2023"

### Supplementary information

#### A. Questionnaires for end-users and stakeholders

##### A.1 - Questionnaire for wastewater treatment plants

**Background information**

1. Which unit/department do you represent? (Fill in)
2. What is your role/responsibilities? (Fill in)

**About future wastewater surveillance and collaboration**

3. How has the collaboration with the laboratory and those who collect the samples worked? Is there anything that could have been done better? (Describe briefly)
4. How has the collaboration with NIPH worked? Is there something that you have missed or that could have been done better? (Describe briefly)
5. What capacity do you have to continue with sampling for this type of surveillance in the future, beyond the test period? (Describe briefly)
6. What will be the biggest challenges for you related to continuing with sampling for this type of purpose? (Describe briefly)

##### A.2 – Questionnaire for public health authorities at local level

**Background information**

1. Which municipality do you represent? (Fill in)
2. Which unit/department do you represent? (Fill in)
3. What is your role? (Fill in)
4. What main tasks and areas of responsibility has your unit/department had in connection with the pandemic? (Fill in)

**Previous knowledge of wastewater surveillance (prior to NIPH pilot)**

5. Do you know whether your municipality has been involved in previous projects where wastewater has been used to map information about the health status of the inhabitants of your municipality? **Yes** ☐ **No** ☐ **Unsure** ☐
6. If yes, what type of mapping? (tick)
   a. Antimicrobial Resistance (AMR) ☐
   b. Other infectious diseases ☐
   c. Chemical substances/medicines/narcotics ☐
   d. Other (please specify) …

**Communication**

7. Which of the following information channels do you use to keep yourself updated on the results of the wastewater surveillance? (tick)
    a. Result report sent by e-mail from NIPH ☐
    b. NIPH’s weekly report (weekly report for COVID-19, influenza, and other respiratory infections ☐
    c. The project’s website (https://www.fhi.no/hn/statistikk/overvaking-smittsomme-sykdommer-i-avlopsvann/) ☐
    d. Other professional actors/channels ☐
8. If you have to choose, through which of the channels mentioned above do you prefer to receive results from the wastewater surveillance? **a. ☐ b. ☐ c. ☐ d. ☐**
9. Which parts of the results did you find most interesting and relevant to the task you are responsible for? (Describe briefly)
10. Do you think the results were understandable and sufficient for the tasks you are responsible for? **Yes☐ No☐** - If no, what do you think could have been done better/differently? (Describe briefly)
11. Is there anything you are missing in the results reports you have received from NIPH? (Describe briefly)
12. Are you satisfied with how frequently you have received results report from NIPH? **Yes, satisfied☐ No, could be more frequent ☐ No, could be less frequent ☐**
13. Are there any other actors/units in your municipality that you think would benefit from being involved in the project and/or receiving results reports? **Yes ☐ No☐ -** If "yes", which actors/units? (Describe briefly)

**Usefulness**

14. In what way have the results been used in your municipality? (Describe briefly)
15. Have the results from the wastewater surveillance been useful to you?

    **Yes ☐ No☐ Unsure ☐ -** If yes, in which way? (Describe briefly)

16. Do you know of any specific measures or assessments that have followed from the results of the wastewater surveillance? **Yes ☐ No☐.**
    a. If "yes”, briefly describe which measures/assessments?
    b. If "no", what do you think is the most important obstacle to the results being able to be used for specific measures or assessments? (Describe briefly)
17. What type of information would be useful to know about the wastewater surveillance besides the information you have received? (Describe briefly)
18. Based on experiences from the test project, do you think that you would have benefited from the results of the project if the wastewater surveillance would have been started earlier in the pandemic, i.e., before June 2022? **Yes ☐ No ☐**

a. If "yes", during which phase of the pandemic do you think you would have benefited the most from wastewater surveillance? **2020 ☐ 2021 ☐ 2022 (prior to June ☐** Please explain why (describe briefly):

**Implementation and cooperation**

19. How did you experience the collaboration with NIPH in connection with the project, do you have any suggestions for something that could have been better? (Describe briefly)

**About future wastewater surveillance**

20. Would you be willing to continue participating in the wastewater surveillance if extended beyond the test period, i.e., after March 2023? **Yes ☐ No ☐ Unsure ☐**
21. Are there any challenges to participating from your side and, if so, what are the biggest obstacles? (Describe briefly)
22. Which other diseases/health threats do you think will be most relevant to include in wastewater-based surveillance in the future, and which will have the highest relevance for you? (Please explain why)

**Suggestion for improvement**

23. Do you/you have other suggestions on how we can improve the system?

#### A.3 – Questionnaire for risk assessors and managers at national level

##### A.3.1 Questions for the Directorate of Health

**Background information**

1. Which unit/department do you represent?
2. What is your role/responsibilities?

**Usefulness and future use**

3. Have you benefited from the results of NIPH's pilot for wastewater surveillance of SARS-CoV-2? In what way? What do you think the results can be used for in the future? (Describe briefly)
4. How do you consider the usefulness of the wastewater surveillance compared to other indicators used in the national monitoring of SARS-CoV-2? (Describe briefly)
5. From a public health perspective, do you have any thoughts about future areas of use for wastewater surveillance? (Describe briefly)
6. What do you think will be the most important prerequisites for wastewater surveillance to be used as a national preparedness tool in dealing with future epidemics and health threats? (Describe briefly)

##### A.3.2 Questions for the Norwegian Institute of Public Health

**Background Information**

1. Which unit/department do you represent?
2. What is your role/responsibilities?

**Usefulness, limitations, and future applications**

3. Have the results of the SARS-CoV-2 wastewater surveillance pilot been useful? In what way? (Describe briefly)
4. Seen from a national surveillance perspective, will there be a need for wastewater surveillance beyond March 2023? **Yes ☐ No ☐ Unsure ☐**
    4.1. If yes, at what level and what do you think the results can be used for in the future? (Describe briefly)
5. Have the results been communicated in an understandable way? Do you have suggestions for improvements in the way we present the results? (Describe briefly)
6. What do you think are the most important limitations of the results from the wastewater surveillance? (Describe briefly)
7. How do you assess the usefulness of the wastewater surveillance compared to other indicators used in the national surveillance of SARS-CoV-2? (Describe briefly)
8. Are there other parts of the surveillance that can be scaled down in the future if we continue with wastewater surveillance? (Describe briefly)
9. If the project had started earlier during the pandemic, do you think the results would have had an impact on NIPH's risk assessments and advice regarding measures? (Describe briefly)
10. From a public health perspective, do you have any thoughts about future areas of use for wastewater surveillance? If the surveillance is to be extended to other agents, which ones do you think should have the highest priority and why? (Describe briefly)
11. What do you think will be the most important prerequisites for wastewater surveillance to be used as a national preparedness tool in addressing future epidemics and health threats? (Describe briefly)

#### B. Supplementary Figures

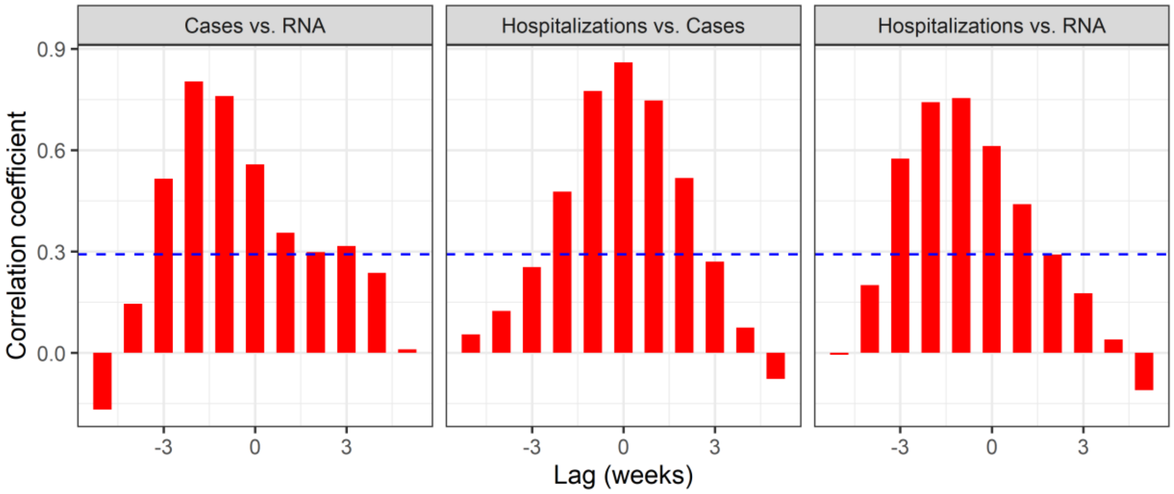


**Figure S1.** Cross correlation between relative daily change in viral load in wastewater (RNA detected in wastewater) and clinical indicators (positive cases and hospitalizations) at various lags (weeks) at increasing trend. Values above the blue dotted line indicate that the correlation is significant ($p \leq 0.05$).

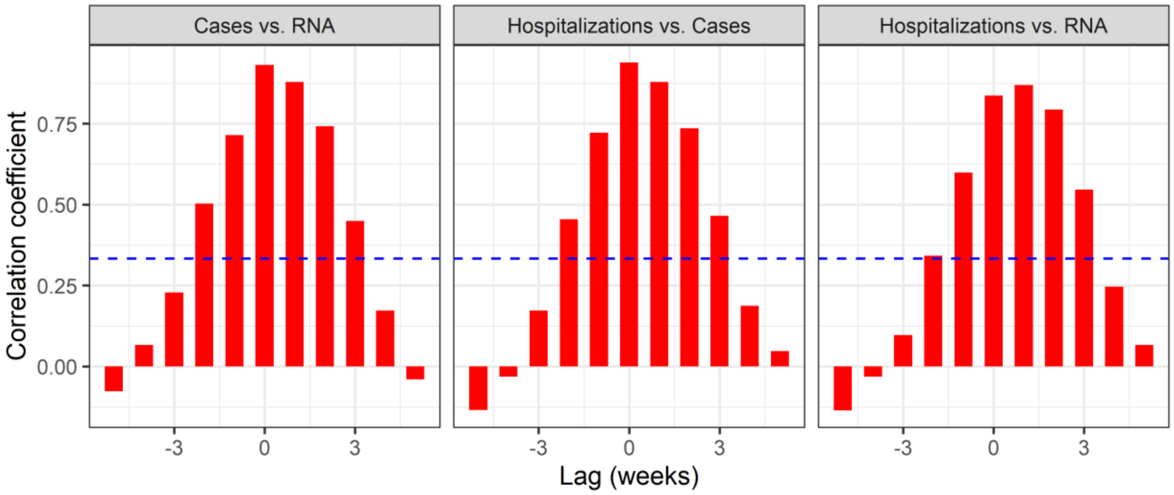


**Figure S2.** Cross correlation between relative daily change in viral load in wastewater (ww RNA) and clinical indicators (hospitalizations and positive cases) at various lags (weeks) at decreasing trend. Values above the blue dotted line indicate that the correlation is significant ($p \leq 0.05$).

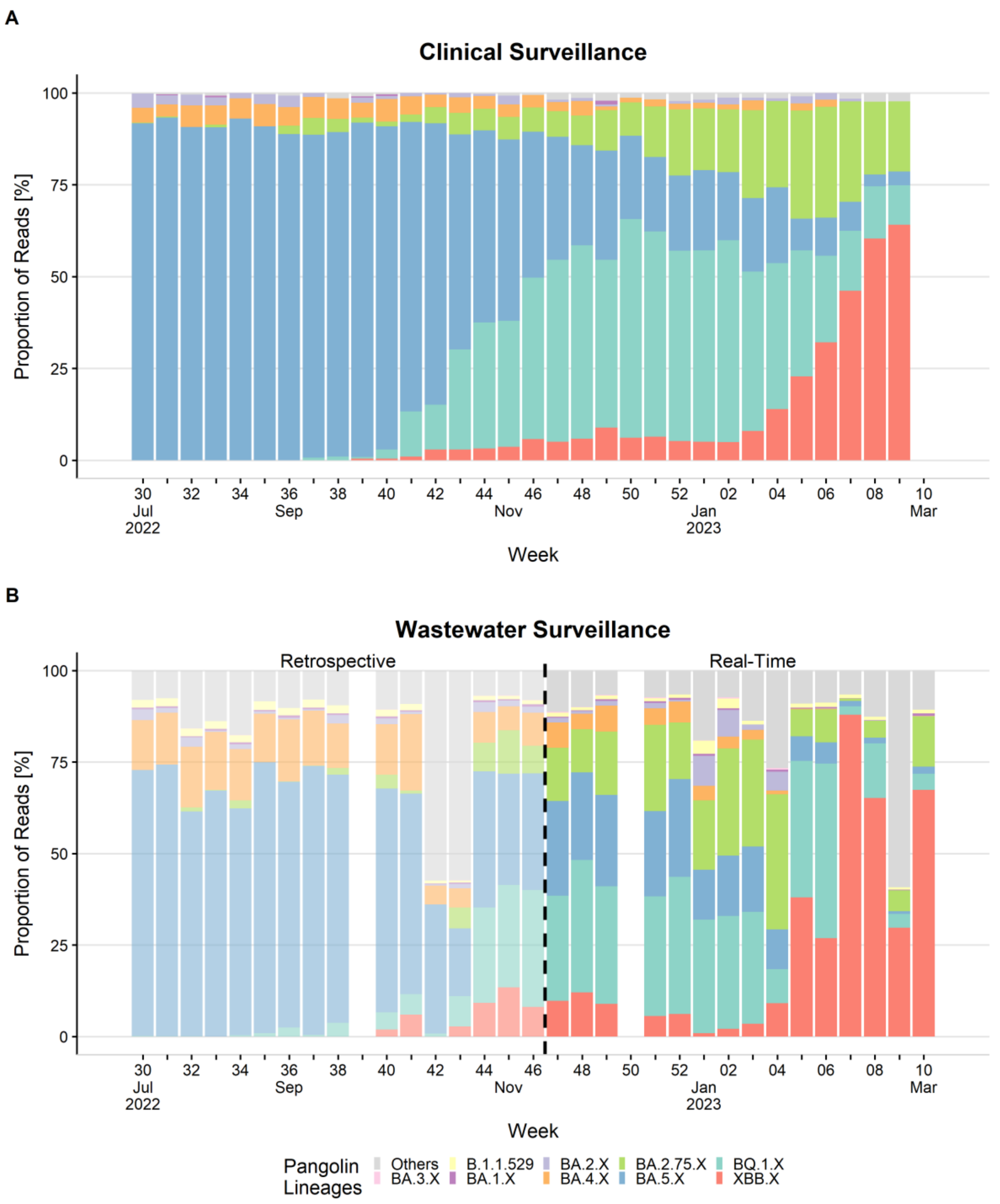


**Figure S3.** Proportion of variants identified in clinical samples for COVID-19 surveillance (A) and wastewater samples including both retrospective and real-time results (B) at national level in Norway. Note: Sequencing results and proportion of variants were available in real-time and reported in the NIPH's weekly reports from week 47 and 5, respectively.
